# Estimating the risk of 2019 Novel Coronavirus death during the course of the outbreak in China, 2020

**DOI:** 10.1101/2020.02.19.20025163

**Authors:** Kenji Mizumoto, Gerardo Chowell

**Author notes:** Address for correspondence: Kenji Mizumoto, Graduate School of Advanced Integrated Studies in Human Survivability, Kyoto University, Yoshida-Nakaadachi-cho, Sakyo-ku, Kyoto 606-8306, Japan.

## Abstract

Since the first case of Novel Coronavirus (2019-nCov) was identified in December 2019 in Wuhan City, China, the number of cases continues to grow across China and multiple cases have been exported to other countries. The cumulative number of reported deaths is at 637 as of February 7, 2020. Here we statistically estimated the time-delay adjusted death risk for Wuhan as well as for China excluding Wuhan to interpret the current severity of the epidemic in China. We found that the latest estimates of the death risk in Wuhan could be as high as 20% in the epicenter of the epidemic whereas we estimate it ∼1% in the relatively mildly-affected areas. Because the elevated death risk estimates are likely associated with a breakdown of the medical/health system, enhanced public health interventions including social distancing and movement restrictions should be effectively implemented to bring the epidemic under control.

## Background

Since the first case of Novel Coronavirus (2019-nCov) was identified in December 2019 in Wuhan City in the Hubei Province of China, the number of cases continues to expand across China, with multiple exported cases reported in over 25 countries. In particular, the cumulative number of deaths is at 637 as of February 7, 2020, a figure that will soon surpass the number of people that succumbed to Severe acute respiratory syndrome (SARS) in 2002-2003 *(1)*.

In the context of an emerging infectious disease with pandemic potential, it is critical to assess not only its ability to spread between humans, but also the associated risk of death from the disease. In particular, the type and intensity of public health interventions are often a function of these epidemiological metrics. Importantly, in the absence of vaccines or antiviral for SARS-CoV and 2019-nCov, the effective implementation of non-pharmaceutical interventions (social distancing) and movement restrictions, which are the only alternative strategies available to mitigate their spread in the population, also pose significant pressure on the global economy *(2)*.

As interventions are gradually implemented and calibrated during the course of an outbreak, early estimates of the case fatality risk (CFR) provide crucial information for policy makers. Yet, the estimation of epidemiological characteristics including the CFR during the course of an outbreak tend to be affected by right censoring and ascertainment bias *(3-5)*. The phenomenon of right censoring is caused by the gap of illness onset to death between vulnerable population and healthy population, resulting in underestimation, while ascertainment bias is due to the unreported bulk of cases with mild symptoms or asymptomatic infection, potentially leading to overestimation.

The purpose of this study is to derive estimates of the time-delay adjusted CFR for 2019-nCov for Wuhan, China as well as for China excluding the city of Wuhan with quantified uncertainty in order to interpret the current severity of the epidemic in China.

## Method

### Data sources

We use two different types of data. First, daily confirmed cases and deaths in China were extracted from daily reports published by China government, Hubei Province in China and

Wuhan City *(7-9)*. These data were categorized by area of Wuhan City, Hubei Province excluding Wuhan City or China excluding Hubei Province, considering that over 50% of the deaths are occurring in Hubei Province. In particular, the great majority of the deaths have occurred in Wuhan City. The diagnosis of 20190-nCov cases relies solely on polymerase chain reaction (PCR), as rapid diagnostic tests for this novel coronavirus are yet to be developed.

Second, fifty epidemiological descriptions of the patients who died of 2019-nCov infection were obtained from several sources (7-9). After checking for duplication and missing data, the sample size of delays from onset to death and from hospitalization to death was 39 and 33, respectively. We fitted a gamma distribution, an exponential distribution and a lognormal distribution to the distributions and selected the best model based on the AIC. (Figure S1-S2, Table S1-S2).

### Case fatality ratio

Crude CFR is defined as the number of cumulative cases divided by the number of cumulative deaths at a specific point in time. For the estimation of CFR in real time, we employ the delay from hospitalization to death, *h*_s_, which is assumed to be given by *h*_s_ = *H*(s) – *H*(s-1) for *s*>0 where *H*(s) follows a gamma distribution with mean 10.1 days and SD 5.4 days, obtained from the available observed data. Let *π* be the time-delay adjusted case fatality ratio, the likelihood function of the estimate *π* is

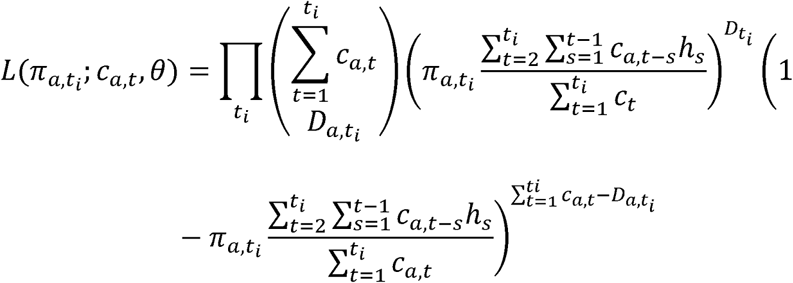

where *c*_*a,t*_ represents the number of new cases with reported day *t* in area *a*, and *D*_*a,t*_ is the number of new deaths with reported day *t*_i_ in area *a* (*10-11*). We assume that the observed cumulative death, *D*_t_ is the result of the binomial sampling process with probability *π*.

We estimated model parameters using a Monte Carlo Markov Chain (MCMC) method in a Bayesian framework. Posterior distributions of the model parameters were estimated by sampling from the three Markov chains. For each chain, we drew 100,000 samples from the posterior distribution after a burn-in of 20,000 iterations. Convergence of MCMC chains were evaluated using the potential scale reduction statistic (*12-13*). Estimates and 95% credibility intervals for these estimates are based on the posterior probability distribution of each parameter and based on the samples drawn from the posterior distributions.

All statistical analyses were conducted in R version 3.6.1 (R Foundation for Statistical Computing, Vienna, Austria) using the ‘rstan’ package.

## Results

As of February 5th, 2020, a total of 28001 cases and 563 deaths of 2019-nCov have been reported in China. Of the 28001 cases reported in China, 11618 cases (37.3%) are from Wuhan City, 10494 cases (33.7%) are from Hubei Province excluding Wuhan City and 8976 cases (28.9%) are from China excluding Hubei Province. Of the 642 deaths in China, 478 deaths (74.5%) are from Wuhan city, 140 deaths (21.9%) are from Hubei Province and only 24 deaths (4%) are from China excluding Hubei Province.

Figure 1 illustrates the cumulative cases in (A) Wuhan City, (B) Hubei Province excluding Wuhan City and (C) China excluding Hubei Province, and the cumulative deaths in (D) Wuhan City, (E) Hubei Province excluding Wuhan City and (F) Mainland China excluding Hubei Province, respectively. The curve of the cumulative number of deaths grows after that of the cumulative number of cases, and the increase in the number of deaths in Wuhan City is occurring more rapidly and the associated mortality burden appears to be much higher relative to rest of China, while the cumulative case counts for the three areas in China are similar.

**Fig 1:**
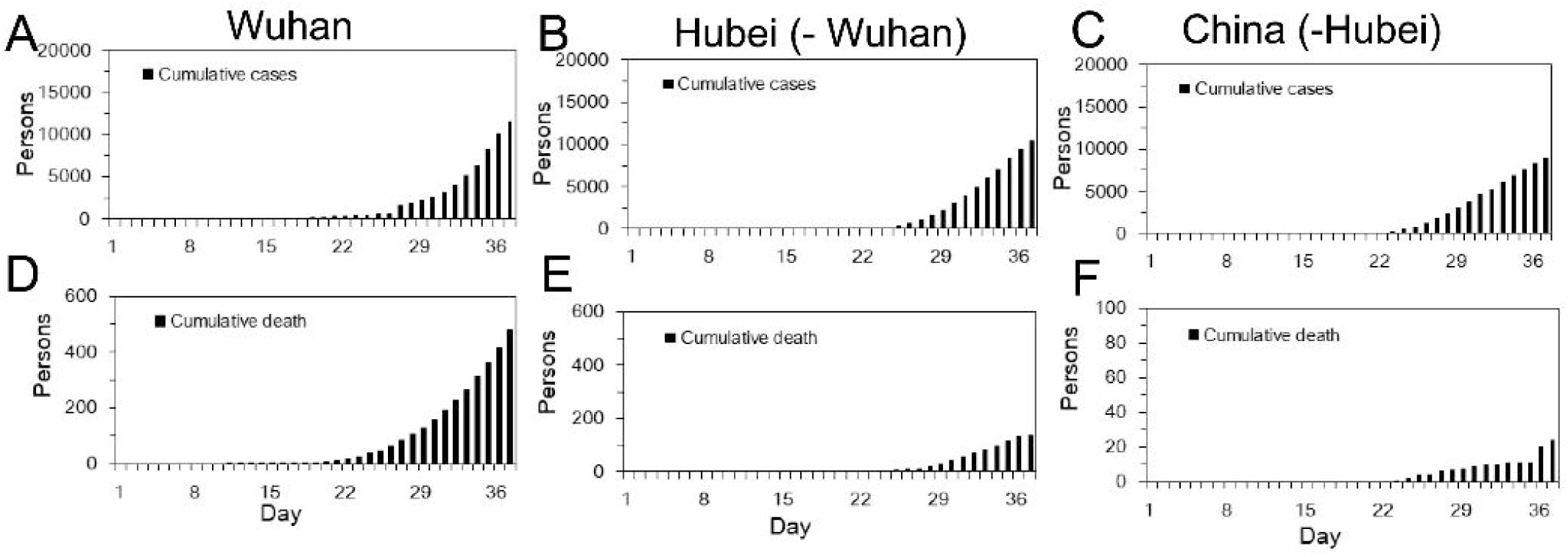
Temporal distribution of cases and deaths by area due to 2019-nCov, China, January – February 2020. Cumulative cases in (A) Wuhan City, (B) Hubei Province excluding Wuhan City and (C) Main land China excluding Hubei Province, and cumulative deaths in (D) Wuhan City, (E) Hubei Province excluding Wuhan City and (F) Main land China excluding Hubei Province, over time. Day 1 corresponds to January 1^st^ in 2020. As the dates of illness onset were not available, we used dates of reporting.

Figure 2 displays the observed and model-based posterior estimates of crude CFR in (A) Wuhan City, (B) Hubei Province excluding Wuhan City and (C) Main land China excluding Hubei Province, and model-based posterior estimates of time-delay adjusted CFR in (D) Wuhan City, (E) Hubei Province excluding Wuhan City and (F) Mainland China excluding Hubei Province, January – February in 2020, respectively. Our model-based crude CFR fitted the observed data well throughout the course of the epidemic except for the very early stage. During the course of the outbreak, our model-based posterior estimates of time-delay adjusted CFR take much higher values than the observed crude CFR, excluding at the early stage in Wuhan City and the later stage in China excluding Hubei Province. Our estimates of the time-delay adjusted CFR appear to be decreasing almost consistently in Hubei Province excluding Wuhan City and in China excluding Hubei Province, whereas in Wuhan city, estimates were low at the early stage and then increased and peaked in the midst of the study period, and are now following a decreasing trend similar to the other two areas, reaching estimates around 20%.

**Fig 2:**
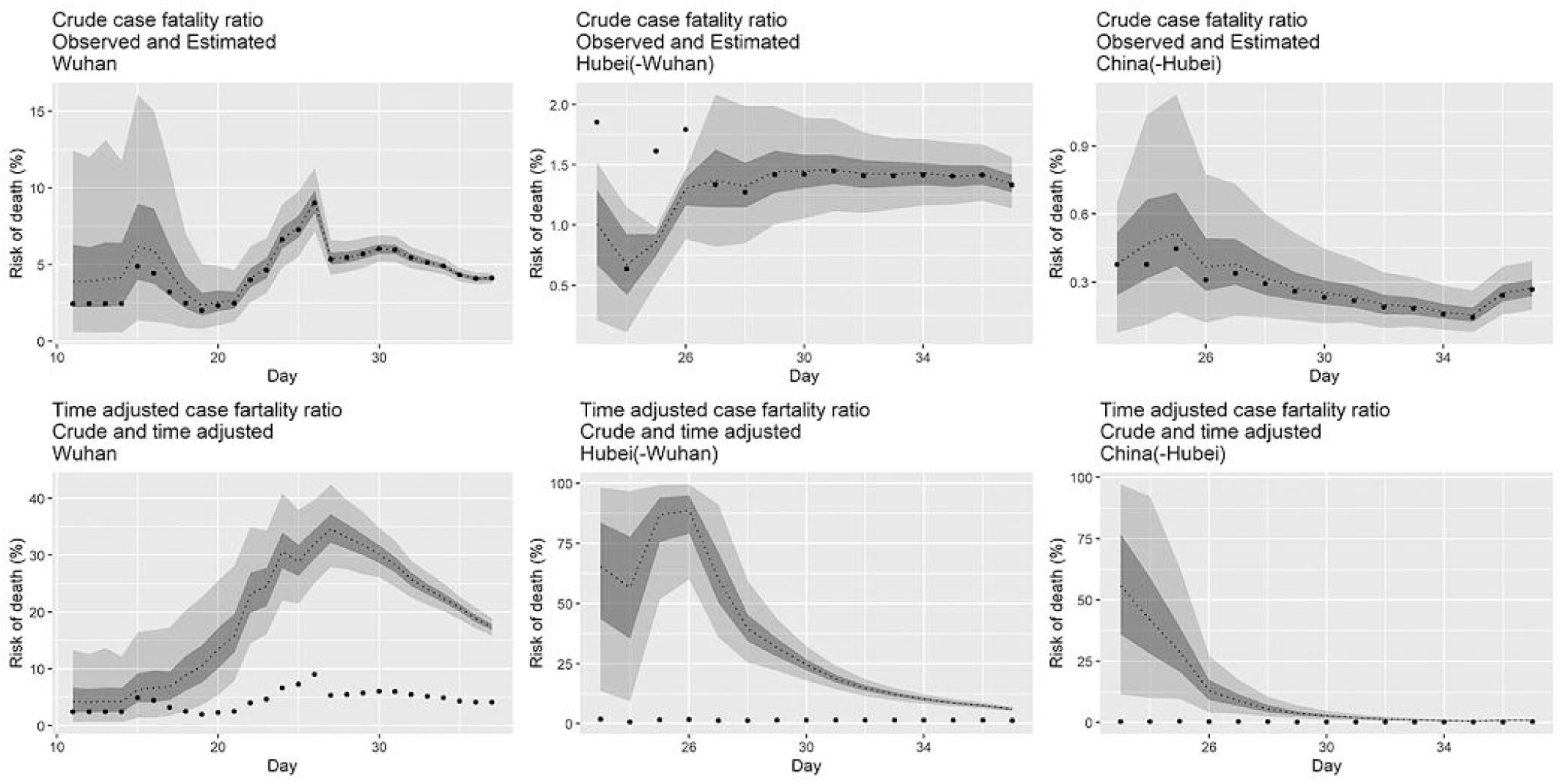
Temporal variation of risk of death caused by 2019-nCov, China, January – February 2020. Observed and posterior estimated of crude case fatality ratio in (A) Wuhan City, (B) Hubei Province excluding Wuhan City and (C) Main land China excluding Hubei Province, and time-delay adjusted case fatality ratio in (D) Wuhan City, (E) Hubei Province excluding Wuhan City and (F) Main land China excluding Hubei Province. Day 1 corresponds to January 1^st^ in 2020. Black dots shows crude case fatality ratio, and light and dark indicates 95% and 50% credible intervals for posterior estimates, respectively.

The latest estimates of the time-delay adjusted CFR are at 18.9% (95%CrI: 17,1-20.8%) in Wuhan City, 6.1% (95%CrI: 5.2-7.1%) in Hubei Province excluding Wuhan City and 0.9% (95%CrI: 0.6-1.3%) in China excluding Hubei Province, while the observed crude CFR takes 4.1% (95% Confidence Interval: 3.8%-4.5%) in Wuhan City, 1.3% (95%CI: 1.1-1.6%) in Hubei Province excluding Wuhan City and 0.27 % (95%CI: 0.2-0.4%) in China excluding Hubei Province, respectively. (Figure 3, Table)

**Table.**
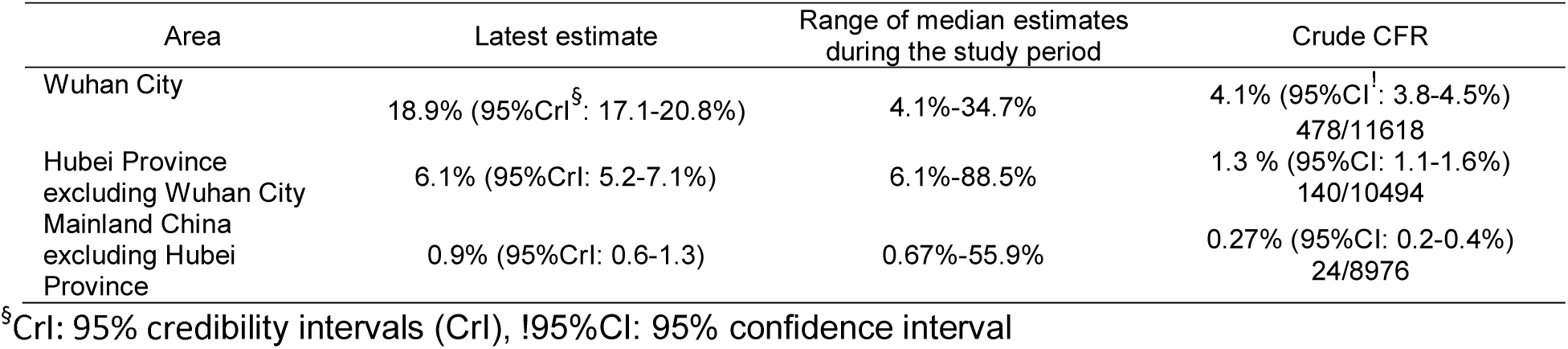
Summary results of time-delay adjusted case fatality ratio of 2019-nCov in the 3 areas in China, 2020 (As of February 6, 2020)

| Area | Latest estimate | Range of median estimates<br>during the study period | Crude CFR |
| --- | --- | --- | --- |
| Wuhan City | 18.9% (95%CrI<sup>§</sup>: 17.1-20.8%) | 4.1%-34.7% | 4.1% (95%CI<sup>!</sup>: 3.8-4.5%)<br>478/11618 |
| Hubei Province<br>excluding Wuhan City | 6.1% (95%CrI: 5.2-7.1%) | 6.1%-88.5% | 1.3 % (95%CI: 1.1-1.6%)<br>140/10494 |
| Mainland China<br>excluding Hubei<br>Province | 0.9% (95%CrI: 0.6-1.3) | 0.67%-55.9% | 0.27% (95%CI: 0.2-0.4%)<br>24/8976 |

<sup>§</sup>CrI: 95% credibility intervals (CrI), !95%CI: 95% confidence interval

**Figure 3.**
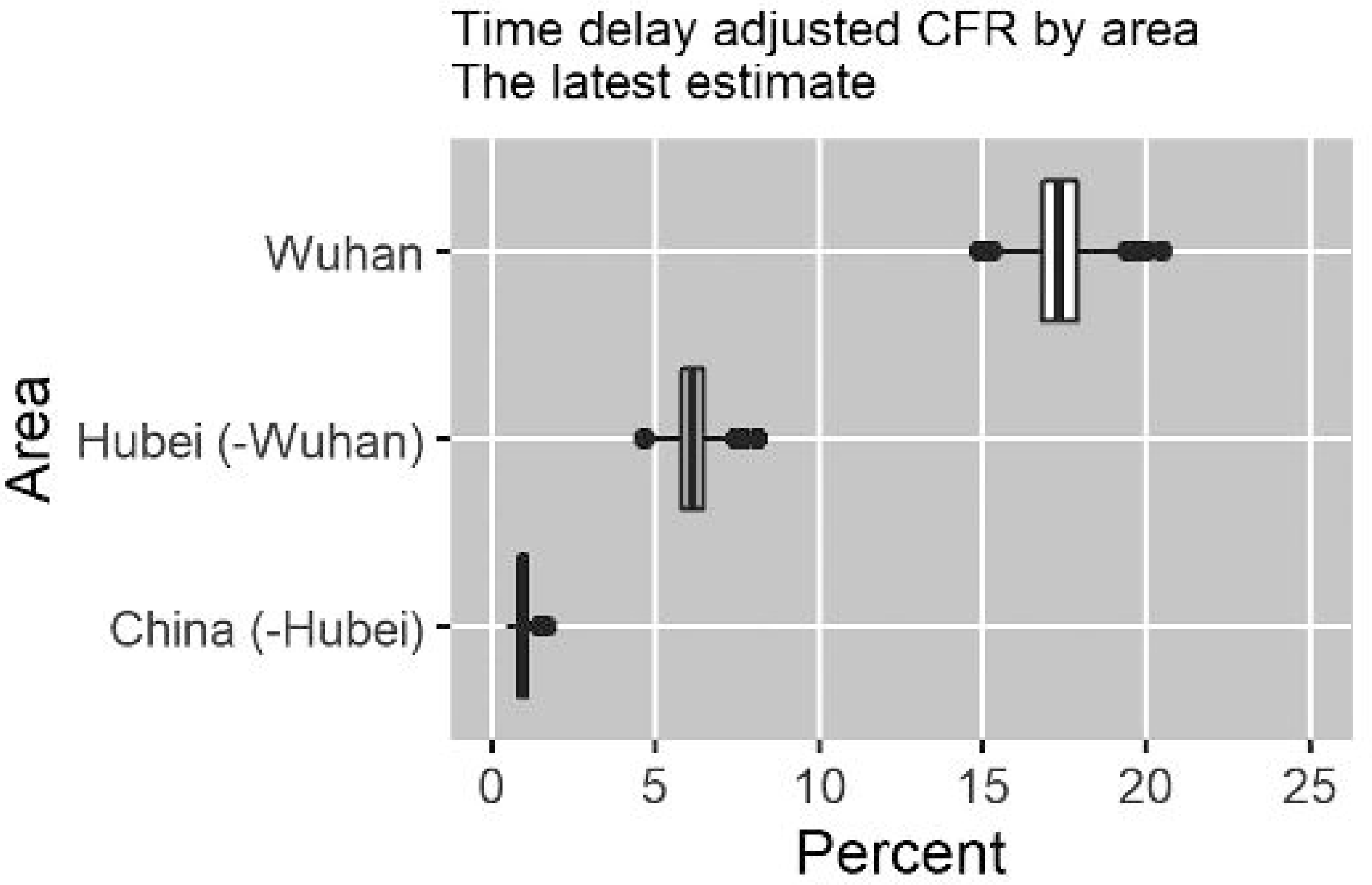
Latest estimates of time-delay adjusted risk of death caused by 2019-nCov by area, 2020, China. Distribution of time-delay adjusted case fatality risks derived from the latest estimates (February 6, 2020) are presented. Top: Wuhan City, Middle: Hubei Province excluding Wuhan City, Bottom: Mainland China excluding Hubei Province.

## Discussion

In this paper we have derived estimates of the CFR for the ongoing 2019-nCov outbreak in China. We have estimated time-delay adjusted CFR in three different areas in China and found that the most severely affected areas are Wuhan City as well as the Hubei Province excluding Wuhan City while the rest of China exhibits a milder impact (China excluding Hubei Province).

We found that the latest estimates of the delay adjusted CFR in Wuhan City is as high as 18.9% (95%CrI: 17.1-20.8%), an estimate that his 3-fold higher than our estimate for Hubei Province excluding Wuhan City and around 20-fold higher than our estimate for China excluding Hubei Province. These findings suggest that the ongoing situation in Wuhan is particularly dire compared to the other affected areas. Upward trend of CFR at the early phase generally indicates a deterioration of an ascertainment bias.

However, this upward trend should be interpreted with caution. In particular, the difficulties in diagnosis cases of 2019-Cov because the associated symptoms are not specific. Further, the fraction of 2019-Cov patients who develop mild symptoms including asymptomatic patients is not minor, which complicates detection and diagnosis early after symptoms onset, leading to ascertainment bias (*14-15*). Indeed, out of a total of 566 Japanese returnees who evacuated Wuhan city by government-chartered plane during January 29-31 in 2020, a total of 5 asymptomatic and 4 symptomatic 2019-nCov cases were detected after detailed medical examination (*16*). However, considering that this underestimation occurred during the course of outbreak and the number of deaths is relatively accurately reported, the upward trend indicates that the temporal disease burden exceeded the capacity of medical facilities and the surveillance system likely missed many cases during the early phase. Additionally, hospital-based transmission is likely occurring, affecting healthcare workers, inpatients and visitors to medical facilities, which could explain an increase trend and elevated CFR estimates. Indeed, nosocomial outbreaks of MERS and SARS nosocomial outbreaks have been reported (*17-18*). Inpatients with underlying disease or seniors infected in the hospital setting have raised the CFR to values as high as 20% for MERS (*19-20*). More generally, 2019-nCov transmission is facilitated in confined settings. For instance, a large cluster of 2019-Cov secondary infection has been reported aboard a cruise ship, with 61 confirmed cases representing about a fifth of the people tested for the virus, supporting high transmissibility of 2019-nCov in confined conditions *(21)*.

A downward trend in CFR is suggestive of the extent of improvements in epidemiological surveillance. In addition, this pattern indirectly supports a significant number of mild or asymptomatic cases in Wuhan City, and the underlying transmission might prolong the end of the outbreak or further transmission to other areas unless effective social distancing measures are implemented while a vaccine becomes available. Furthermore, given that both the delay adjusted CFR and crude CFR estimates in Wuhan City are around 20-fold higher than our estimates for China excluding Hubei Province suggests a breakdown in medical care delivery and points to the critical need for urgent medical support in the epicenter of the epidemic.

We also found that the estimates of delay adjusted CFR both in in Hubei Province excluding Wuhan City and in China excluding Hubei Province show a time-dependent declining trend as the epidemic progresses. Indeed, a similar trend has been previously reported for the Middle East Respiratory Syndrome (MERS) outbreak in the Republic of Korea in 2015, where a significant fraction of the cases had comorbidities or were senior patients (*16-17*). If the proportion of vulnerable cases is high at the early phase and gets proportionally smaller in the later stage of the outbreak, this partly explains the time dependent decline. However, since the epidemic has yet to peak, this time dependent decrease is likely caused by ascertainment bias.

Moreover, both of the latest estimates of the delay adjusted CFR and crude CFR in Hubei Province are around 6-7 fold higher than that in China excluding Hubei Province, where the health care system has not reached peak capacity. This also indicates the need to anticipate for additional medical support to deliver medical care to the most vulnerable and at highest risk of succumbing to the disease from underlying medical conditions.

Our study is not free from limitations. First, our CFR estimate is influenced by ascertainment bias and this might influence estimates upwards. For those infectious diseases characterized by a large fraction cases with mild or asymptomatic illness, the infection fatality risk, e.g., the number of death divided by the total infected, is a more appropriate index of disease burden (*22-23*). For this purpose, mass serological surveillance and surveys to assess the presence/absence of symptoms is strongly recommended to disentangle the threat of emerging infectious diseases including 2019-nCov.

Second, in our estimation we employed the time delay distribution from illness onset to deaths (N=39), which was obtained from secondary sources, but the available data does not include either the date of illness onset or the confirmed date. For this reason, we utilized the time delay from hospitalization to death (N=33).

In conclusion, our latest estimates of the risk of 2019-Cov deaths in China could be as high as 20% in the epicenter of the epidemic whereas this estimate is around 1% in the relatively mildly-affected areas in China as of February 5^th^, 2020. Because it is likely that the death risk from 2019-nCov is associated with a breakdown of the medical/health system in the absence of pharmaceutical interventions (vaccination and antiviral drugs), enhanced public health interventions including social distancing, quarantine, enhanced infection control in healthcare settings and movement restrictions should be effectively implemented to rapidly contain this epidemic.

## Data Availability

The present study relies on published data and access information to essential components of the data are available from the corresponding author.

## Acknowledgments

KM acknowledges support from the Japan Society for the Promotion of Science (JSPS) KAKENHI Grant Number 15K20936, from Program for Advancing Strategic International Networks to Accelerate the Circulation of Talented Researchers Grant Number G2801 and from the Leading Initiative for Excellent Young Researchers from the Ministry of Education, Culture, Sport, Science & Technology of Japan.

GC acknowledges support from NSF grant 1414374 as part of the joint NSF-NIH-USDA Ecology and Evolution of Infectious Diseases program; UK Biotechnology and Biological Sciences Research Council grant BB/M008894/1.

**Appendix Table S1. Summary of delay distribution from hospitalization to death, 2020, China**.

Gamma, exponential and exponential distribution were fitted to the available observed data, and estimated parameters and AIC for each distribution are presented (N=33).

**Appendix Table S2. Summary of delay distribution from illness onset to death, 2020, China**.

Gamma, exponential and exponential distribution were fitted to the available observed data, and estimated parameters and AIC for each distribution are presented (N=39).

**Appendix Figure S1. Delay distribution from hospitalization to death, 2020, China**.

Gamma (dark, dashed), exponential distribution (light black, dashed) and kernel density distribution (black, full) were fitted to the available observed data (N=33).

**Appendix Figure S2. Delay distribution from illness onset to death, 2020, China**.

Gamma (dark, dashed), exponential distribution (light black, dashed) and kernel density distribution (black, full) were fitted to the available observed data (N=39).

